# Based on body temperature, CRP, Platelet and Age Scoring Models in distinguishing sterile inflammatory fever and infectious fever in patients with acute Myocardial Infarction

**DOI:** 10.64898/2025.12.25.25343029

**Authors:** Jingshu Lei, Junxian Song, Qinghao Zhao, Lina wang, Kuibao Li, Ruiying Wang, Wei Zhao

## Abstract

**Background:** Fever is a common clinical manifestation following acute myocardial infarction(AMI). However, differentiating its etiology-specifically, distinguishing between fever caused by the absorption of necrotic tissue and fever due to concurrent infection is critical, as this distinction directly influences treatment decisions and patient prognosis.

**Methods:** This retrospective study reviewed AMI patients from two centers. Participants were divided into two groups: a fever of non-infectious origin group and patients with fever secondary to infection.We initially identified candidate risk factors demonstrating significant differences, Then,a stepwise selection technique was applied to potential univariate correlates (P < 0.1) to construct a multivariable risk score model.

**Results:** 69 out of 1507 AMI patients were included, with 32 patients (46.4%) presenting AMI complicated by infection and 37 patients (53.6%) having AMI without infection but presenting with fever.Twelve dichotomous characteristics were screened as candidate predictor variables. Using stepwise elimination, four independent predictors were retained in the final logistic model: age, peak body temperature (BT), baseline platelet count, and peak CRP. Multivariate analysis identified the following independent predictors:age (OR=1.11, 95% CI: 1.041-1.186), peak BT (OR=9.83, 95% CI: 1.954-49.441), baseline platelet count (OR=1.01, 95% CI: 0.999-1.029), and peak CRP (OR=1.01, 95% CI: 0.998-1.026).A risk score was calculated by assigning points to categorized levels of each predictor weighted by their respective regression coefficients. The scoring distribution were: age (0,1,2,3,4,5 points), peak BT (0, 1, 3, 5 points), baseline platelet count (0, 1, 3, 5, 6 points), and peak CRP (0, 1, 2 points), which ranged from a minimum of 0 to a maximum of 18 .The score could predict the risk of infection with good discriminative ability (AuROC: 0.899, 95% CI: 0.823-0.975 P=0.000) in fever of AMI patients.

**Conclusions:** We first demonstrate that a simple bedside score incorporating BT, CRP, platelet count, and age can effectively discriminate between sterile inflammatory fever and infectious fever in patients with AMI.

## Introduction

Acute myocardial infarction (AMI) is a critical condition, and co-infection is not uncommon among its patients, particularly the elderly. Reported hospital-acquired infection rates range from 14.8% to 27%,which could prolong hospital stays, elevate higher healthcare costs, and increased mortality.Fever and elevated inflammatory markers typically indicate infection; However, in AMI patients, they can also be caused by myocardial cell necrosis.Differentiating between an infectious fever and non-infectious systemic inflammatory response in AMI patients is often challenging, as both scenarios are associated with elevated inflammatory markers. This situation carries the dual risks of exacerbating antibiotic resistance through over-treatment and catastrophic consequences from under-diagnosis. The differentiation between sterile inflammation and co-infection in AMI is complicated by the absence of well-defined, time-dependent thresholds for inflammatory markers, a critical knowledge gap that current limited research has yet to address.Accordingly, this study was designed to develop a novel risk stratification score that is both simpler and more practical, with the objective of predicting infection and distinguishing the cause of fever in AMI patients.

## METHODS

### Selection of Participants

This research was based on a retrospective review of the clinical data from AMI patients who were admitted to two tertiary hospitals: Peking University People’s Hospital and Hebei Yanda Hospital.To ensure the enrollment of an unbiased population, all discharged patients with a confirmed diagnosis of AMI, were retrospectively enrolled. The inclusion criterion was applied to 1507 consecutive patients identified through the electronic medical record system between January 1, 2022, and April 30, 2024.

Patient enrollment criteria were as follows: Patients were classified as the case group if they had a confirmed AMI diagnosis along with fever and a subsequent diagnosis of infection. Those with AMI and fever but without any confirmed infection were assigned to the control group. To rigorously ensure the absence of infectious factors in the control group, the inclusion criteria were set very strictly.Firstly,the patient reported none infection-related symptoms;Secondly, clinical examination was negative for signs of infection, including: no wet rales or wheezing on lung auscultation; no pharyngeal congestion or edema,etc;Third, all microbiological cultures obtained within 48 hours of admission were negative. Additionally, respiratory viral nucleic acid amplification tests, including for influenza and SARS-CoV-2, returned negative results;Fourthly, no radiological evidence of infection was found across all modalities;Fifth, the patient had not received any anti-infective agents at any point, and no empirical therapy for a suspected infection was initiated during the hospitalization;Sixth, no major infectious complications—defined as sepsis, septic shock, or multiple organ dysfunction syndrome (MODS)—occurred during hospitalization included at least 7 days of clinical observation.

Patients were excluded if the available clinical data were inadequate to definitively determine the presence or absence of a concurrent infection. Pediatric patients (age < 18 years) were also excluded.Additional exclusion criteria comprised: Other significant cardiac conditions (e.g., rheumatic heart disease, dilated cardiomyopathy, congenital heart disease);Any comorbidities known to confound inflammatory markers, specifically including stress-induced cardiomyopathy and myocarditis; severe hepatic dysfunction (Child-Pugh class C); significant renal impairment (CKD stage 4-5 or acute kidney injury); advanced malignancies; major hematologic diseases; or active autoimmune disorders.

#### Data Collection and Definition

Data extracted from the electronic medical records of AMI patients included a comprehensive array of clinical and demographic information, along with hospital outcomes.Laboratory markers of inflammation comprised admission and peak body temperature (BT), complete blood count parameters at baseline and peak (white blood cell, neutrophil, lymphocyte, and platelet counts; platelet distribution width and volume), and baseline and peak C-reactive protein (CRP) levels.

Standardized definitions were applied to all patient variables and clinical diagnoses.AMI was defined according to the Fourth Universal Definition of Myocardial Infarction, and STEMI was defined by the 2023 ESC guidelines^[1]^.Fever was considered present if the axillary temperature was ≥37.1°C^[2]^.The diagnosis of pulmonary infection^[3]^, urinary system infection^[4]^, upper respiratory tract infection^[5]^ and COVID-19 infection^[6]^ was based on their corresponding established diagnostic criteria.

### Statistical analysis

We employed clinical and laboratory variables in analysis to identify predictors for the study endpoints.Continuous variables with a normal distribution are expressed as mean ± standard deviation (SD), whereas those with a non-normal distribution as median with interquartile range (P25, P75). Categorical variables were presented as frequencies and percentages (%), and compared using the chi-square test or Fisher’s exact test, as appropriate.First, the differential variables between two groups was used to select relevant predictors. Then, stepwise multivariate logistic regression was employed to identify independent risk factors.Statistical analyses were conducted with SPSS 16.0 (IBM Corp., Armonk, NY, USA). Statistical significance was set at P < 0.05.

### Model Development and Score derivation

The primary endpoint was defined as the infection either at admission or during hospitalization.Variables with significant differences between groups were initially selected as candidate risk factors for further analysis.The stepwise logistic regression incorporated demographic and clinical variables along with laboratory values,variables with a univariate association of P < 0.1 were entered to derive the final multivariable model.A risk score was assigned to each final predictor based on its logistic regression coefficient. Specifically, points were derived from the magnitude of the β-coefficients.

### Statement of ethics

We retrospectively extracted all patient data from medical records and maintained them in a dedicated research archive.The study was conducted in accordance with the Declaration of Helsinki and was approved by the local ethics committee of Heibei Yanda Hospital.

## RESULTS

### Patient population characteristics

The final study population consisted of 69 patients selected from 1,507 with AMI based on strict inclusion criteria; 75.4% (n=52) were male, and the mean age was 69.0 ± 15.3 years (range: 30–94).Of the 69 patients, 32 (46.4%) presented with AMI complicated by infection and fever, and 37 (53.6%) had AMI with fever but no infection. STEMI was diagnosed in 41 patients (59.4%) and non-STEMI in 28 (40.6%).Co-infections predominantly involved: pulmonary (30.4%), urinary tract (7.2%), upper respiratory tract (5.8%), COVID-19 (10.1%), and other sites (7.2%).The baseline clinical characteristics are summarized in Table 1.

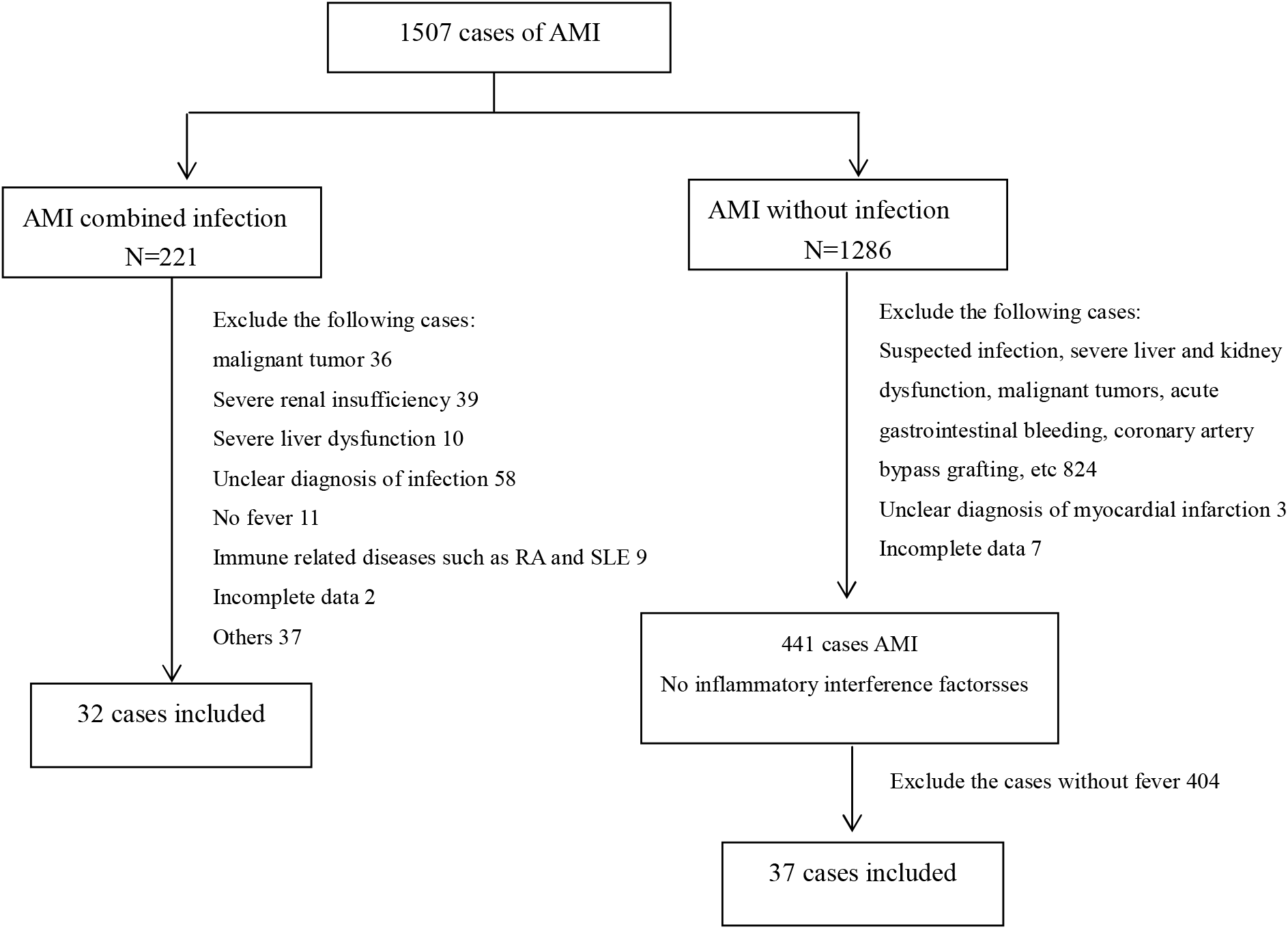

**Table 1.** showed the clinical characteristics of the global patient population.

| Clinical characteristics | Results |
| --- | --- |
| Age | 69.03±15.3 |
| Male ( n,%) | 52(75.4%) |
| STEMI | 41(59.4%) |
| Non-STEMI | 28(40.6%) |
| Combined fever without infection | 37(53.6%) |
| Coinfection | 32(46.4%) |
| lung infection | 21(30.4%) |
| urinary tract infection | 5(7.2%) |
| upper respiratory tract infection | 4(5.8%) |
| NCP infection | 7(10.1%) |
| Other infection | 5(7.2%) |

Baseline comorbidities were largely balanced between the two groups. Compared to patients without infection, those with infection were older (69.97 ± 13.74 vs. 57.03 ± 14.10 years, p < 0.001), had a higher prevalence of diabetes (59.4% *vs*. 24.3%, p = 0.003), and exhibited higher peak body temperature (38.17 ± 0.67 *vs*. 37.57 ± 0.43°C, p < 0.001), longer fever duration (3.75 ± 2.65 *vs*. 1.95 ± 1.25 days, p = 0.001), and higher platelet distribution width both at baseline (14.32 ± 2.83 *vs*. 12.90 ± 2.39 fL, p = 0.027) and peak (15.40 ± 2.51 *vs*. 14.21 ± 2.58 fL, p = 0.058).Peak CRP was markedly higher in the infection group [50.65(13.90,144.88) *vs*. 7.65(3.27,58.45), P = 0.003]. In contrast, both baseline lymphocyte count (1.57 ± 1.12 *vs*. 2.28 ± 1.21, P = 0.014) and peak lymphocyte count (1.36 ± 0.87 *vs*. 1.79 ± 0.77, P = 0.033) were significantly lower.The incidence of in-hospital major cardiovascular events [cardiopulmonary resuscitation, malignant arrhythmia, heart failure(HR), death] differed between groups. Notably, HR occurred more frequently in patients with infection (34.4% *vs*. 5.4%, p = 0.002) (Table 2).

**Table 2.** Displays the clinical characteristics of both groups.

| Variables | Coinfection<br>32(46.4) | Absorb heat<br>37(53.6) | P |
| --- | --- | --- | --- |
| Age | 69.97±13.74 | 57.03±14.10 | 0.000 |
| Diabetes | 19(59.4) | 9(24.3) | 0.003 |
| Hypertension | 23(71.9) | 22(59.5) | 0.280 |
| Cerebral infarction | 10(31.3) | 5(13.5) | 0.075 |
| Smoking | 15(46.9) | 23(62.2) | 0.203 |
| Drinking history | 8(25.0) | 14(37.8) | 0.254 |
| Malignant arrhythmia | 8(25.0) | 8(21.6) | 0.704 |
| Death | 1(3.1) | 1(2.7) | 0.917 |
| Cardiopulmonary resuscitation | 2(6.3) | 3(8.1) | 0.767 |
| Heart failure | 11(34.4) | 2(5.4) | 0.002 |
| Baseline body temperature | 36.56±0.55 | 36.52±0.30 | 0.677 |
| Peak body temperature | 38.17±0.67 | 37.57±0.43 | 0.000 |
| Fever duration | 3.75±2.65 | 1.95±1.25 | 0.001 |
| Baseline WBC | 11.48±4.31 | 10.58±4.03 | 0.374 |
| Baseline neutrophil | 11.08±10.93 | 9.36±11.85 | 0.534 |
| Baseline lymphocyte | 1.57±1.12 | 2.28±1.21 | 0.014 |
| Baseline platelet | 232.50(160.75,289.50) | 216.00(184.50,265.00) | 0.904 |
| Baseline platelet distribution width | 14.32±2.83 | 12.90±2.39 | 0.027 |
| Baseline platelet distribution volume | 9.26±2.54 | 10.07±0.98 | 0.076 |
| Baseline CRP | 5.79(2.20,73.78) | 2.45(0.90,23.28) | 0.071 |
| Peak WBC | 11.16±4.02 | 9.86±2.91 | 0.129 |
| Peak neutrophil | 8.77±3.75 | 8.83±9.69 | 0.977 |
| Peak lymphocyte | 1.36±0.87 | 1.79±0.77 | 0.033 |
| Peak platelet | 186.00(153.25,265.75) | 216.00(175.50,259.00) | 0.149 |
| Peak platelet distribution width | 15.40±2.51 | 14.21±2.58 | 0.058 |
| Peak platelet distribution volume | 9.36±2.65 | 9.81±1.15 | 0.355 |
| Peak CRP | 50.65(13.90,144.88) | 7.65(3.27,58.45) | 0.003 |

Twelve dichotomous variables were screened as candidate predictors. These included age, STEMI diagnosis, diabetes, HF, peak BT, along with first lymphocyte count, and platelet count, platelet distribution width, and CRP levels at both baseline and peak (derived from Tables 1 & 2). These candidate variables were then following stepwise logistic regression (entry criterion: P < P<0.1), four independent predictors were retained in the final model for infection risk algorithm: age, peak BT, baseline platelet count, and peak CRP.

The main risk factors were incorporated into a multivariate logistic regression to derive their regression coefficients (β), odds ratios (ORs), and corresponding 95% confidence intervals (CIs).The results were presented in Table 3. Age (OR = 1.11, 95% CI: 1.04–1.19) and peak BT (OR = 9.83, 95% CI: 1.95–49.44) were statistically significant predictors. Baseline platelet count (OR = 1.01, 95% CI: 1.00–1.03) and peak CRP (OR = 1.01, 95% CI: 1.00–1.03) were retained in the model but did not reach statistical significance,but the matching became the best model combination.

**Table 3.** Best multivariable clinical predictors for the final logistic model.

|  | P | OR | 95% CI |
| --- | --- | --- | --- |
| Age | 0.001 | 1.11 | 1.041-1.186 |
| Peak body temperature | 0.006 | 9.83 | 1.954-49.441 |
| First platelet | 0.061 | 1.01 | 0.999-1.029 |
| Peak CRP | 0.086 | 1.01 | 0.998-1.026 |

OR: odds ratio;CI: confidence interval;;CRP:C-reactive protein

#### Risk score

A risk score was calculated by assigning points to categorized levels of each predictor, weighted by their respective regression coefficients. The scoring distribution were: age (0–5 points); peak BT (0, 1, 3, 5 points); baseline platelet count (0, 1, 3, 5, 6 points); and peak CRP (0, 1, 2 points), as specified in Table 4.

**Table 4.**
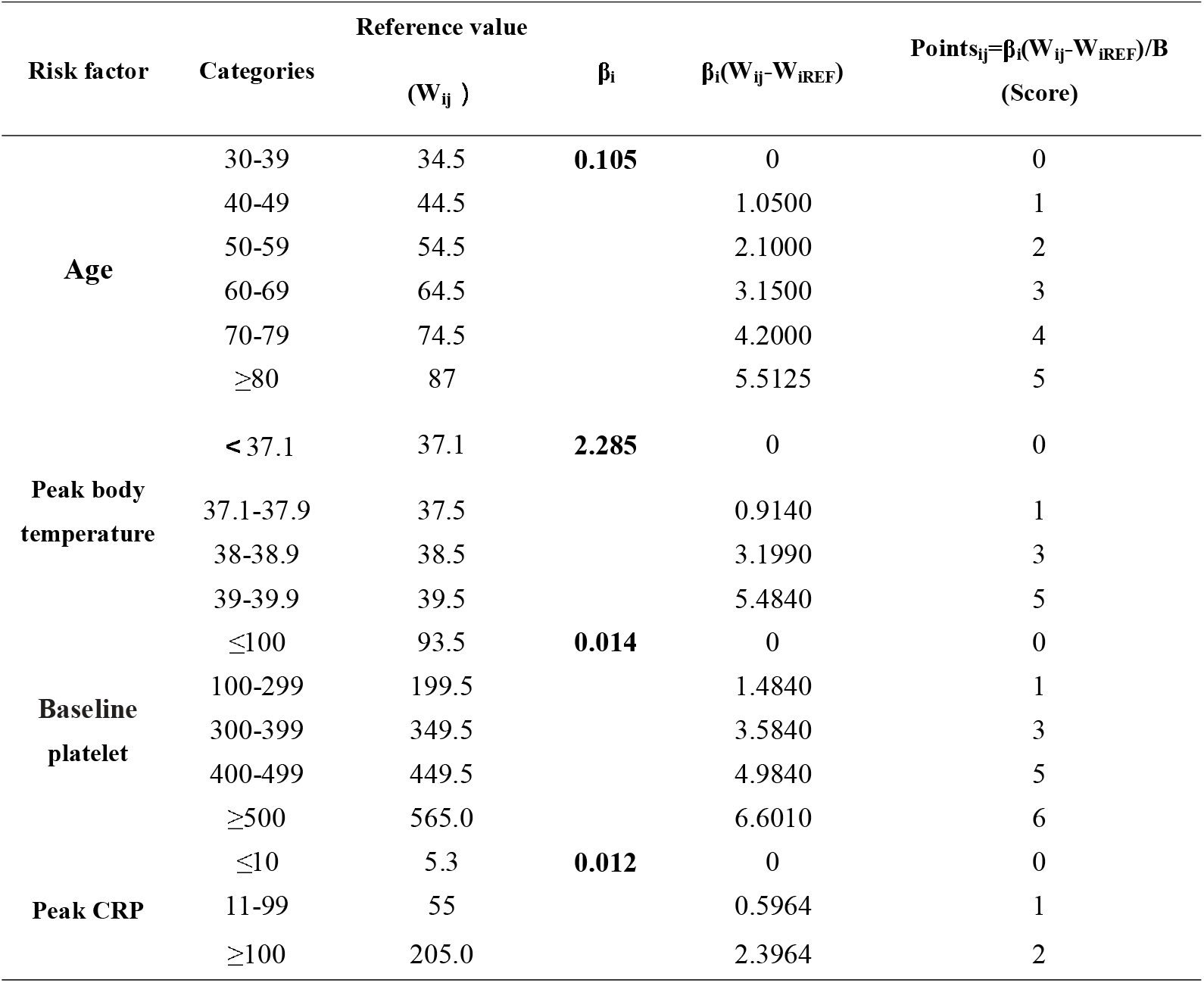
Risk Prediction Model and Point score for each risk factors.

#### Risk Prediction Model

The newly developed risk stratification tool was termed the Predictive Model for Infection in Patients with Acute Myocardial Infarction (PMI-AMI) Score, with a range of 0 to 18 score (Table 4).The predictive infection risk with good discriminative ability (Table 5), with a corresponding predicted probability ranging from 0.2% to 99%.

**Table 5.** Point Score Risk Prediction.

| Total Points | Estimate of risk,% | Total Points | Estimate of risk,% |
| --- | --- | --- | --- |
| 0 | 0.00065227 | 10 | 0.959519024 |
| 1 | 0.001861703 | 11 | 0.985451328 |
| 2 | 0.005301763 | 12 | 0.994860261 |
| 3 | 0.015002829 | 13 | 0.995109704 |
| 4 | 0.041710381 | 14 | 0.999367754 |
| 5 | 0.110622332 | 15 | 0.999778662 |
| 6 | 0.26223194 | 16 | 0.999922535 |
| 7 | 0.503899921 | 17 | 0.999972891 |
| 8 | 0.743759257 | 18 | 0.999990513 |
| 9 | 0.892410196 |  |  |

Based on these predicted probabilities, patients were classified into five progressive risk categories, corresponding to event rates of 0%, 11%, 50%, 89%, and ≥95%. We classified patients into low- and high-risk groups based on PMI-AMIS score cut-off of 7: scores < 7 indicated low risk, while scores ≥ 7 indicated high risk (Table 5).

The PMI-AMI score demonstrated good discriminative ability in predicting infection, with an area under the receiver operating characteristic curve (AuROC) of 0.899 (95% CI: 0.823–0.975, P < 0.001) (Figure 1). The corresponding sensitivity and specificity were 0.875 and 0.833, respectively. Furthermore, the Hosmer-Lemeshow test indicated good model fit ( P = 0.835).

**Figure 1.**
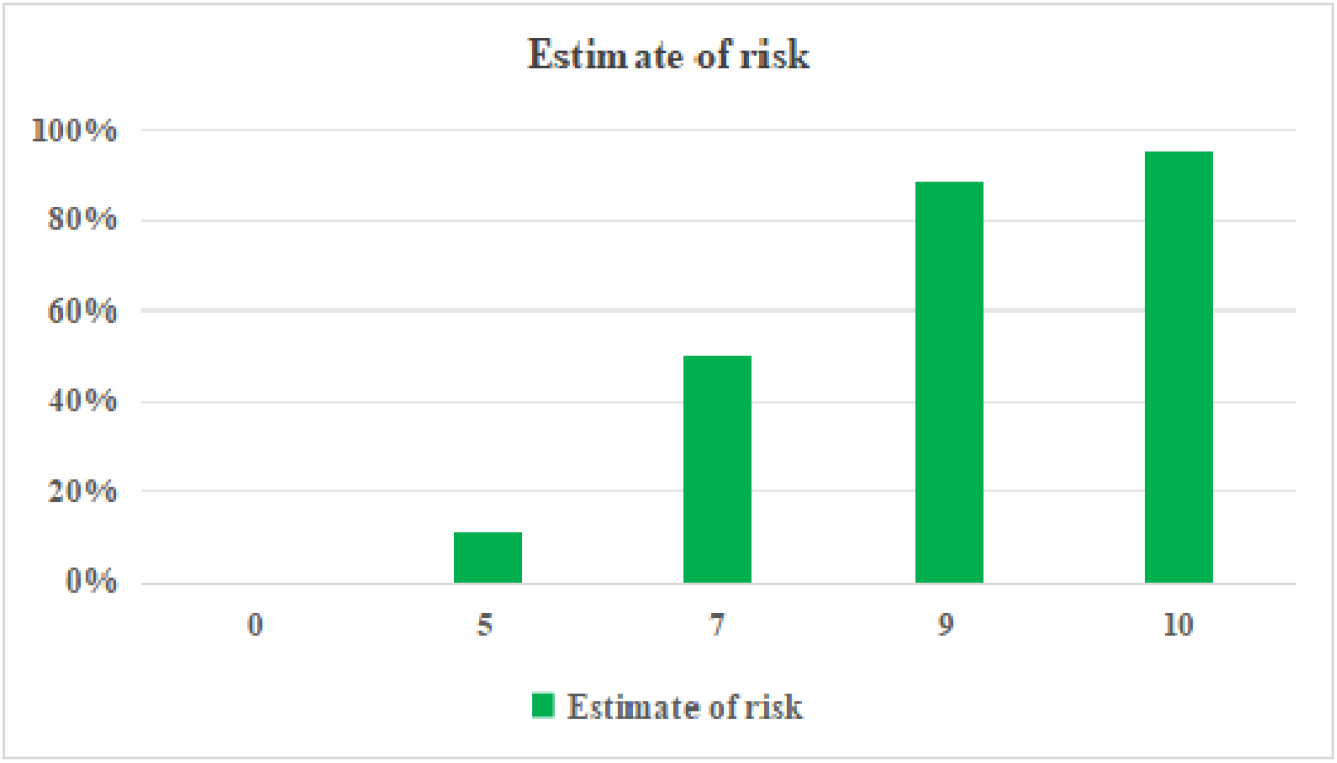
Risk categories according to risk score: very low-risk (0 points, primary end point 0%), low-risk (5 point, primary end point11%), intermediate-risk (7 points, primary end point 50%), high-risk (9 points, primary end poin t89%), and very high-risk (≥10 points, primary end point ≥95%).

**Figure 2.**
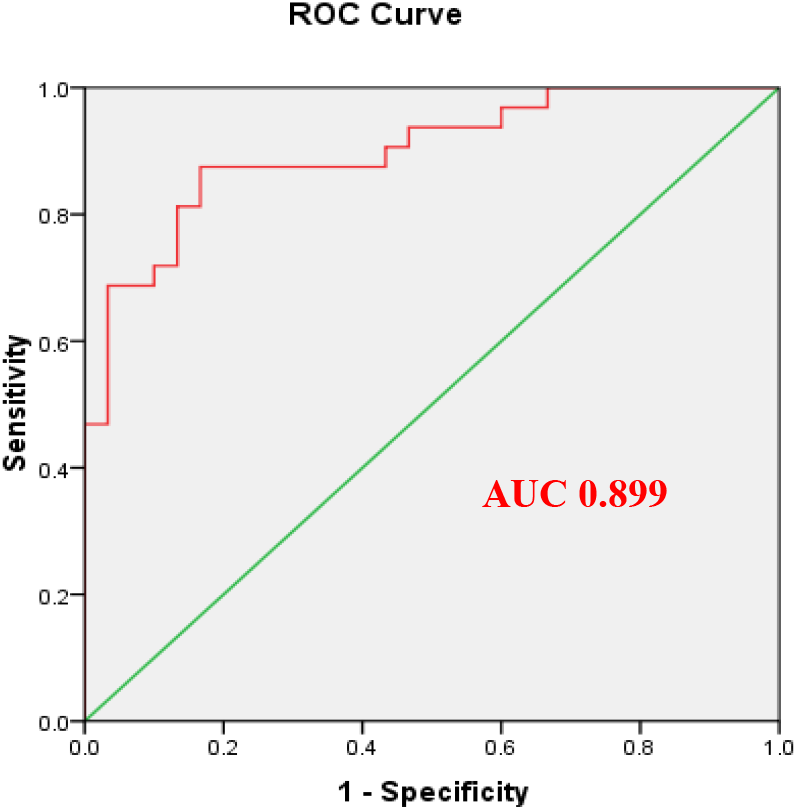
Receiver operating curve demonstrating the Predictive Model for Infection in Patients with Acute Myocardial Infarction (PMI-AMI) score to predict infection among AMI patients. Area under the curve was 0.899 demonstrating excellent discrimination.

## DISCUSSION

The differentiation of fever etiology in AMI patients—whether due to myocardial necrosis or concurrent infection—is a critical clinical scenario that directly determines treatment strategy and influences prognosis.Our study developed a simple scoring model that leverages commonly available clinical indicators—BT, CRP, platelet count, and patient age—all of which are routinely assessed in practice.The model exhibited robust performance in discriminating between the two fever etiologies, with an AuROC of 0.899.

CRP, which as an acute-phase protein is elevated in both infection and tissue injury, was a key discriminator in this model. However, the magnitude and kinetics of its rise may differ significantly between these two conditions.Given that bacterial infection is a powerful inflammatory stimulus, it can drive CRP levels to markedly high values, frequently surpassing 100 mg/L^[7]^. Although myocardial infarction itself, as a sterile inflammatory process, elevates CRP, its peak level and kinetic profile are typically distinct from those observed in severe infections. CRP typically peaks 24-72 hours after symptom onset and serves as a biomarker of the acute sterile inflammatory response triggered by MI^[8].^CRP associated with myocardial tissue repair follows a more protracted course, rising slowly to a plateau before eventually decreasing.Following multivariable adjustment, the model identified significantly elevated CRP levels as associated with infection.The key point was that a markedly elevated CRP often signals a concurrent infection, even in the context of the expected inflammatory response to AMI.While no universal cutoff exists, CRP levels in bacterial infection commonly exceed ten fold the 2 mg/L reference—a threshold often cited for suggesting infectious etiology^[9]^.Researchers have incorporated CRP, a recognized marker of inflammation and cardiovascular risk, into multiple risk stratification models, including the SYNTAX score (SXscore) in ACS^[10]^ and the Reynolds Risk Score^[11]^,ect.

Platelets play a central role in both the initiation and progression of atherosclerosis.The dynamic changes in platelet count offer valuable clues for identification, as platelets influence AMI progression by promoting local thrombosis and mediating inflammation. This is often linked to heightened platelet activation and reactivity^[14]^.In line with our findings, platelet indices were a well-established parameter in AMI research.For instance,by adding PDW and MPV to the GROUP score, HE’s study developed a tool that predicts in-hospital mortality and long-term MACEs in PCI-treated STEMI patients^[17]^.Meta-analysis that involving 18 cohort studies and 16,545 AMI patients, found that higher platelet-to-lymphocyte ratio (PLR) was associated with mortality and MACE^[18]^.

In STEMI patients, fever commonly arises from the systemic inflammatory burden triggered by myocardial tissue necrosis.As an early signal of systemic inflammation, BT elevation is a valuable noninvasive measure for daily clinical use, given that it typically occurs before the increase in CRP^[19]^.The pattern and peak of fever offer differential diagnostic value. Absorptive fever typically presents as a low to moderate elevation (<38.5□), emerging 2-3 days post-infarction with a regular pattern ^[20]^,peaking at approximately 38 hours (±22 hours)^[21]^.Elevated maximum BT was linked to larger infarct size and myocardial injury^[22]^,along with left ventricular remodeling and increased re-hospitalization^[21]^. Infectious fever typically manifests as high-grade (>39°C), remittent, or irregular fever; its onset is often temporally dissociated from the infarction.Although absorptive fever is typically low-grade, our data suggest that the peak BT in infectious fever may be not only higher but also more specific to its etiological type.

Age is an independent risk factor that predisposes individuals to both a higher susceptibility to and greater severity of infection.Due to immune dysfunction, elderly AMI patients are at higher risk for occult infections and may exhibit atypical clinical presentations.The inflammatory response following infection may be blunted in its initial presentation but ultimately more severe.Older patients demonstrated a differential systemic inflammatory profile compared to younger counterparts, characterized by elevated IL-6 and sTNF-RI levels within 48 hours, a lower leukocyte, neutrophil, and lymphocyte counts, and a higher neutrophil-to-lymphocyte ratio (NLR) at 24 hours^[23]^.Multivariate analysis identified age as an independent predictor of febrile etiology, indicating a higher risk of infection. By incorporating age as a variable, the model adjusts for this potential confounder.

This model faces a common clinical dilemma: excessive antibiotic use in febrile AMI patients exacerbates antimicrobial resistance, while failing to identify a genuine infection can lead to catastrophic outcomes. The primary innovation of our study lies in the development of a novel, integrated predictive model that translates four conventional laboratory indicators into a simple bedside score,representing a significant advance in point-of-care diagnostic strategy.This provides an objective, reproducible, and cost-effective decision aid that is simple and rapid to implement at the bedside.This inherent efficiency makes it immediately viable and highly suitable for widespread adoption across all levels of hospitals, particularly in resource-constrained settings such as emergency departments and cardiology wards.This makes our study more akin to real-world research.

### Study limitations

The primary limitations of this study were its retrospective design and the limited sample size, which was largely attributable to our stringent inclusion criteria Our model was deliberately designed around parameters universally available at admission, which necessarily excluded other informative but less universally rapid markers such as procalcitonin (PCT) – known for its specificity in bacterial infections – as well as the dynamic trends of white blood cell counts and interleukins. Future studies should investigate the incremental value of incorporating more specific biomarkers.In addition, the patient’s comorbidities (such as immune suppression status), infarct size, etc. may also affect the cause of fever, which can be considered in future model optimization.It is important to note that the scoring model developed in this study requires external validation.

### Conclusions

In summary, we demonstrate that a simple bedside score incorporating body temperature, CRP, platelet count, and age,which could effectively discriminate between sterile inflammatory fever and infectious fever in patients with AMI.Despite these limitations, this study provides a practical and immediately applicable decision aid for a common clinical dilemma. Prospective studies are now warranted to confirm its efficacy and generalizability, and to explore pathways for its further optimization.

## Statement of Ethics

The study was approved by Medical Ethics Committee of Hebei Yanda Hospital (ID :2021-6-002), and all patients provided written informed consent.

## Author Contributions

Lina wang,Kuibao Li,Jingshu Lei,Qinghao Zhao,Junxian Song designed the research study. Lina wang, Qinghao Zhao, Wei Zhao and Jingshu Lei analyzed the data. Lina wang, Junxian Song, Ruiying Wang and Jingshu Lei draft the manuscript; Jingshu Lei andJunxian Song revise.All authors contributed to editorial changes in the manuscript. All authors read and approved the final manuscript. All authors have participated sufficiently in the work and agreed to be accountable for all aspects of the work.All authors listed have contributed sufficiently to the project to be included as authors, and all those who are qualified to be authors are listed in the author byline.

## Data Availability Statement

The data used during this study are available from the corresponding author on reasonable request.

